# Cerebrospinal Fluid Lactate as a Prognostic Marker of Disease Severity and Mortality in Cryptococcal Meningitis

**DOI:** 10.1101/2020.08.24.20181289

**Authors:** Mahsa Abassi, Ananta S Bangdiwala, Edwin Nuwagira, Kiiza Kandole Tadeo, Michael Okirwoth, Darlisha A Williams, Edward Mpoza, Lillian Tugume, Kenneth Ssebambulidde, Kathy Huppler Hullsiek, Abdu K Musubire, Conrad Muzoora, Joshua Rhein, David B Meya, David R Boulware, ASTRO-CM team

## Abstract

**Background:** Cerebrospinal fluid (CSF) lactate levels can differentiate between bacterial and viral meningitis. We measured CSF lactate in individuals with cryptococcal meningitis to determine its clinical significance.

**Methods:** We measured point-of-care CSF lactate at the bedside of 319 HIV-infected Ugandan adults at diagnosis of cryptococcal meningitis. We summarized demographic variables and clinical characteristics by CSF lactate tertiles. We evaluated the association of CSF lactate with clinical characteristics and survival.

**Results:** Individuals with high CSF lactate >5 mmol/L at cryptococcal diagnosis more likely presented with altered mental status (p<.0001), seizures (p=.0005), elevated intracranial opening pressure (p=.03), higher CSF white cells (p=0.007), and lower CSF glucose (p=.0003) compared to those with mid-range (3.1 to 5 mmol/L) or low (≤3 mmol/L) CSF lactate levels. Two-week mortality was higher among individuals with high baseline CSF lactate >5 mmol/L (35%; 38/109) as compared to individuals with mid-range (22%; 25/112) or low CSF lactate (9%; 9/97; p=<.0001). After multivariate adjustment, CSF lactate >5mmol/L remained independently associated with excess mortality (adjusted Hazard Ratio = 3.41; 95%CI, 1.55-7.51; p=.002). We found no correlation between baseline CSF lactate levels and blood capillary lactate levels (p=.72).

**Conclusions:** Baseline point-of-care CSF lactate levels may be utilized as a prognostic marker of disease severity and mortality in cryptococcal meningitis. Individuals with an elevated baseline CSF lactate are more likely to present with altered mental status, seizures, elevated CSF opening pressures, and are at a greater risk of death. Future studies are needed to determine targeted therapeutic management strategy in persons with high CSF lactate.

**Summary:** In HIV-associated cryptococcal meningitis, baseline cerebrospinal fluid (CSF) lactate levels are associated with increased intracranial pressures, seizures, and altered mental status. Elevated CSF lactate levels, at baseline, are associated with increased 2-week mortality.

## Introduction

The measurement of CSF lactate has been shown to have utility in the rapid differentiation of bacterial meningitis from viral meningitis.[1, 2] In cases of acute meningitis, CSF lactate concentrations of >4.2 mmol/L have a sensitivity of 96% in the presumptive diagnosis of bacterial meningitis.[1] However, despite the high sensitivity, CSF lactate measurements are highly nonspecific and are also found to be elevated in several other infectious and noninfectious neurological conditions, including cerebral malaria, tuberculous meningitis, cryptococcal meningitis, cerebral injury, subarachnoid hemorrhage, seizures, and ischemia.[3-7] As a diagnostic tool, the current recommended clinical application to use CSF lactate, as dictated by the Infectious Diseases Society of America guidelines, is in the diagnostic work up of bacterial meningitis after postoperative neurological procedures.[5, 8]

As a prognostic marker in cases of bacterial meningitis, individuals with higher CSF lactate levels demonstrate a trend towards lower Glasgow coma score and higher mortality.[1, 2] In cases of bacterial meningitis, serial measurements of CSF lactate predict future clinical outcomes. Specifically, a greater than 50% decline in CSF lactate measurements in the first 3 days of treatment resulted in an improved clinical course.[1] A similar trend has been reported in cerebral malaria and in cases of severe head injury, where the normalization of CSF lactate has favorable clinical outcomes, while persistently elevated or increasing CSF lactate levels result in overall poor clinical outcomes including severe disability or death.[3, 5]

In sub-Saharan Africa, HIV-associated cryptococcal meningitis is the most common cause of meningitis, accounting for 15% of AIDS-related deaths globally.[9-11] In low-income countries where the burden of cryptococcal meningitis is greatest, 2-week mortality ranges between 17% and 28% in clinical trials, with one-year mortality as high as 70% in routine care. [9] In HIV-associated cryptococcal meningitis, CSF lactate levels have been found to be elevated (median 3.0 mmol/L (IQR 2.7 to 5.8)) at the time of diagnosis.[4] However, the significance of elevated CSF lactate levels in cryptococcal meningitis is not well understood. We sought to understand the prognostic significance of elevated baseline CSF lactate levels in HIV-associated cryptococcal meningitis.

## Methods

### Study design and participants

The study included Ugandan adults presenting with first-episode cryptococcal meningitis at Kiruddu General Hospital in Kampala and Mbarara Regional Referral Hospital from September 2016 to March 2020. Persons diagnosed with cryptococcal meningitis were enrolled into the Adjunctive Sertraline for the Treatment of HIV-Associated Cryptococcal Meningitis (ASTRO-CM) clinical trial (ClinicalTrials.gov identifier: NCT01802385) through May 2017. After conclusion of ASTRO-CM, participants were enrolled in an observational cohort focused on meningitis diagnostics.[12-14] All participants enrolled into the studies provided written informed consent. Participants were treated with standard antifungal therapy and those who were enrolled into ASTRO-CM were randomized to either receive adjunctive sertraline (400 mg/day for 14 days, followed by 200 mg/day for 12 weeks) or placebo. Standard antifungal therapy was amphotericin B (0.7-1.0 mg/kg/day) for up to 14 days and fluconazole 800 mg/day for ~4 weeks, followed by fluconazole 400 mg/day for 8 weeks of consolidation therapy, and fluconazole 200 mg/day for secondary prophylaxis. Participants in the randomized trial were followed up for 18 weeks, while participants in the observational study were followed for 2 weeks or until hospital discharge.

### Lactate Measurements

All participants had a baseline lumbar puncture performed at the time of cryptococcal meningitis diagnosis. CSF lactate measurements were performed bedside at the time of lumbar puncture (Lactate Plus Analyzer; Sports Resource Group Inc., Hawthorne, NY). In addition to CSF lactate measurements, a subset of individuals also had matched serum lactate measurements performed concurrently at baseline.

### Statistical Methods

CSF lactate measurements were grouped into tertiles. We summarized baseline demographic variables and clinical characteristics by CSF lactate tertiles, presented as percentages and medians with interquartile range. We evaluated the relationship between CSF lactate level and 2-week mortality using log-rank tests and visually summarized by Kaplan-Meier curves. We used Cox proportional hazards models to estimate hazard ratios by CSF lactate level, adjusted for Glasgow coma score <15, baseline seizures, quantitative *Cryptococcus* CSF culture, and CSF opening pressure.

We conducted an area under the receiver operating characteristic (ROC) curve analysis of the continuous log_2_ transformed CSF lactate as a predictor of 2-week mortality to consider a threshold cut point of lactate risk for mortality. As sensitivity analyses, we used Cox proportional hazards models to consider lactate cut at the threshold from the AUC result, and as a continuous variable. We compared clinical characteristics of individuals who died as compared to those who survived at 2-weeks using Cox proportional hazards model. We used the Spearman correlation to estimate the correlation between CSF lactate and serum lactate. We performed analyses using SAS version 9.4 (SAS Institute, Cary, North Carolina).

## Results

Of 622 participants with a first episode of cryptococcal meningitis, 51% (319/622) had a CSF lactate measurement performed at baseline. Demographic and clinical characteristics of participants with a CSF lactate measurement are summarized by CSF lactate tertiles in **Table 1**. CSF lactate measurements (overall median 3.9, IQR [2.8, 5.7], range [1.1, 12.0]) were grouped into low (≤3 mmol/L; n=97), mid-range (3.1 – 5.0 mmol/L, n=113), and high (>5.0, n=109) tertiles with distribution displayed in **Figure 1**.

**Table 1.** Baseline Characteristics of Study Participants by CSF Lactate Tertile.

| Characteristic | N with data | CSF Lactate Tertile |  |  | P-value |
| --- | --- | --- | --- | --- | --- |
|  |  | Low<br>≤3.0 | Mid-Range<br>3.1 to 5.0 | High<br>> 5.0 |  |
| N | 319 | 97 | 113 | 109 |  |
| CSF lactate, mmol/L | 319 | 2.4 (2.1, 2.7) | 3.8 (3.4, 4.4) | 6.7 (5.6, 8.1) |  |
| Men | 319 | 51 (52.6%) | 75 (66.4%) | 79 (72.5%) | 0.01 |
| Age, years | 319 | 34 (29, 40) | 35 (30, 42) | 38 (31, 45) | 0.02 |
| Glasgow Coma Score < 15 | 314 | 30 (31.3%) | 39 (35.5%) | 71 (65.7%) | <0.0001 |
| Seizure | 314 | 9 (9.4%) | 17 (15.5%) | 32 (29.6%) | 0.0006 |
| On HIV therapy | 317 | 50 (52.1%) | 51 (45.1%) | 53 (49.1%) | 0.60 |
| Hemoglobin, g/dL | 239 | 11 (10, 13) | 12 (10, 14) | 11 (9, 13) | 0.29 |
| CD4 cells/μL | 225 | 14 (7, 38) | 15 (6, 53) | 25 (8, 58) | 0.18 |
| Creatinine, mg/dL | 236 | 0.7 (0.6, 0.9) | 0.7 (0.6, 0.9) | 0.8 (0.6, 1.0) | 0.28 |
| CSF white cells/μL | 313 | 4 (4, 20) | 4 (4, 30) | 4 (4, 130) | 0.007 |
| CSF Protein, mg/dL | 270 | 50 (24, 102) | 59 (29, 100) | 65 (28, 122) | 0.25 |
| CSF Glucose, mg/dL | 240 | 62 (42, 86) | 59 (40, 72) | 38 (22, 68) | 0.0003 |
| CSF Opening Pressures, cm H<sub>2</sub>O | 280 | 22 (14, 30) | 23 (16, 33) | 27 (17, 40) | 0.03 |
| CSF culture, log<sub>10</sub> CFU/mL | 306 | 4.0 (1.7, 5.3) | 4.6 (2.5, 5.6) | 4.6 (2.8, 5.4) | 0.25 |
| CSF removed, mL | 290 | 10 (7, 16) | 12 (9, 18) | 12 (8, 19) | 0.22 |
| Blood Capillary Lactate, mmol/L | 37 | 1.9 (1.9, 5.3) | 2.9 (2.0, 4.5) | 3.5 (1.8, 5.0) | 0.72 |

Data are presented as median (IQR) or N (%).
P-value from Kruskal-Wallis test for medians, Chi-squared test for proportions

**Figure 1:**
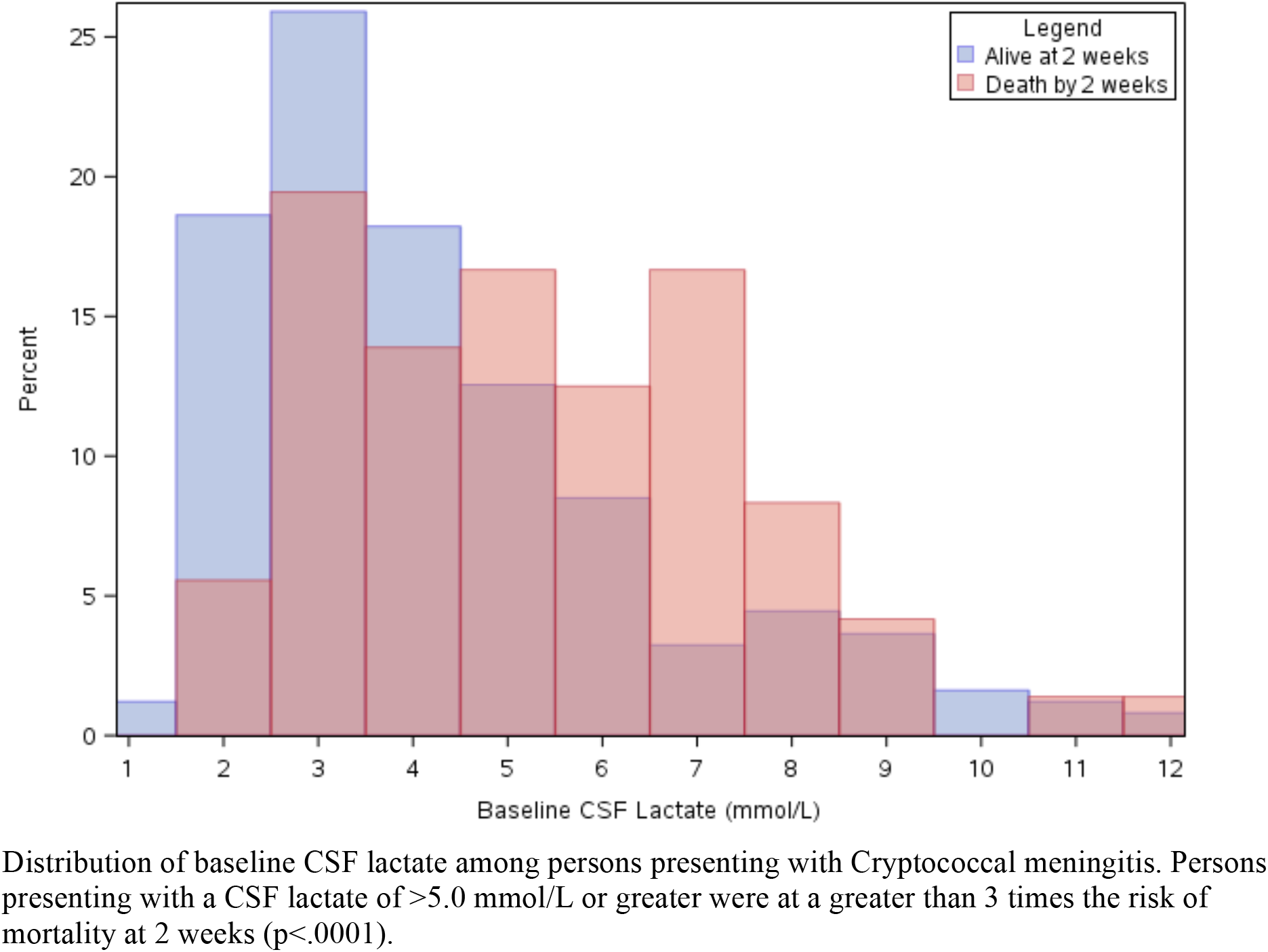
Distribution of Baseline CSF Lactate Measurements by 2-week survival.

Among persons diagnosed with HIV-associated cryptococcal meningitis, we found several demographic and clinical factors to be associated with elevated CSF lactate levels. Participants with high CSF lactate, >5.0 mmol/L, had significantly higher baseline CSF white cells (p=.007) and lower CSF glucose (p=.0003) than participants with low or mid-range CSF lactate measurements at baseline. There were no differences in CSF quantitative *Cryptococcus* culture (p=.25) or CSF protein (p=.25) between lactate tertiles. However, participants with high baseline CSF lactate also had higher intracranial opening pressures (median 27 cm H_2_O IQR, 17 to 40) than participants with mid-range (median 23 cm H_2_O IQR, 16 to 33) or low baseline CSF lactate levels (median 22 cm H_2_O IQR, 14 to 30) (p=.03). A significantly greater percentage of participants with high baseline CSF lactate presented with a Glasgow coma score <15 (71%) as compared to mid-range CSF lactate (39%) or participants with low (30%) (p<.0001). Baseline seizures were also more frequent in participants presenting with high baseline CSF lactate at the time cryptococcal meningitis diagnosis (high: 30% vs. mid-range: 16% and low: 9%; p=.0006).

Two-week mortality was significantly higher among individuals with high baseline CSF lactate compared to individuals with mid-range and low CSF lactate levels (high: 35% (38/109) vs. mid-range: 22% (25/112) and low: 9% (9/97); p<.0001) **(Figure 2)**. In multivariate analysis, after adjusting for Glasgow coma score, baseline seizure, baseline opening pressures, and quantitative *Cryptococcus* CSF culture, individuals with high baseline CSF lactate continued to be at a three-fold higher risk of mortality at 2-weeks compared to those with CSF lactate ≤3.0 mmol/L (adjusted Hazard Ratio = 3.41; 95% CI, 1.55 to 7.51; p=.0021) **(Table 2)**. The increased risk of mortality with higher lactate levels was also seen in sensitivity analysis when considering baseline CSF lactate as continuous variable (aHR=1.13 for each 1 mmol/L increase in baseline CSF lactate, 95% CI, 1.02 to 1.24) **(Table 2)**. An area under a receiver-operator characteristic curve analysis found a CSF lactate cut-off point of 4.3 mmol/L as having the highest sensitivity and specificity for predicting 2-week mortality (AUC: 0.664; 95%CI, 0.60 to 0.73). In sensitivity analysis, those with baseline CSF lactate of 4.3 mmol/L or greater had a two-fold higher risk of mortality at 2 weeks compared to those with baseline CSF lactate < 4.3 mmol/L (aHR 2.10, 95% CI, 1.22 to 3.62) **(Table 2)**.

**Figure 2:**
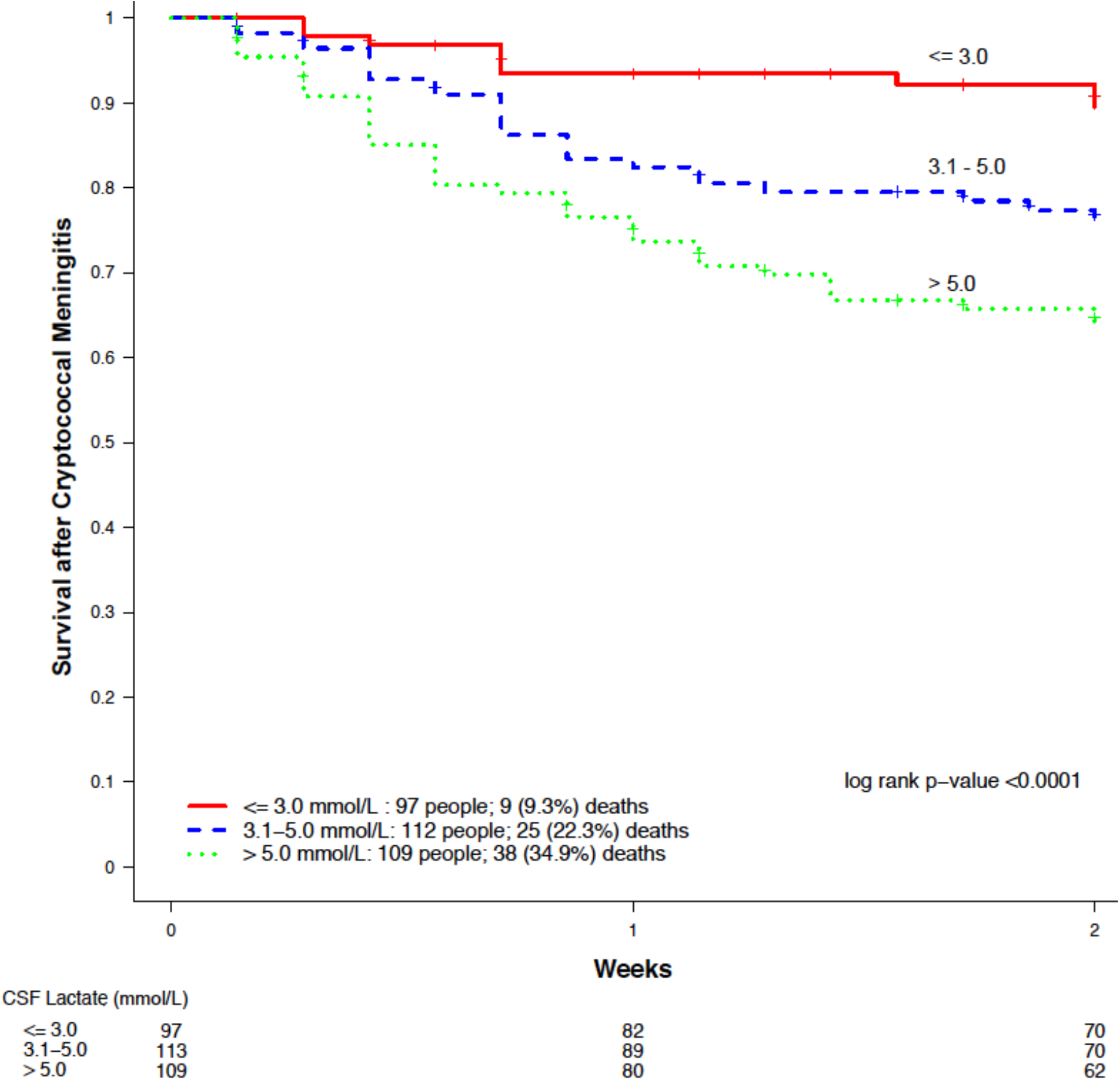
Mortality at 2-weeks by Baseline CSF Lactate Tertiles.

**Table 2:** Hazard Ratios for 2-Week Mortality.

| CSF Lactate<br>(mmol/L) | Deaths<br>N (%) | Hazard Ratio<br>(95%CI) | P-value | Adjusted*<br>Hazard Ratio<br>(95%CI) | P-value |
| --- | --- | --- | --- | --- | --- |
| **By CSF Lactate Tertiles** |  |  |  |  |  |
| **≤3.0** | 9 (9%) | 1.0 |  | 1.0 |  |
| **3.1 to 5.0** | 25 (22%) | 2.51 (1.17, 5.38) | 0.02 | 1.99 (0.87, 4.52) | 0.10 |
| **> 5.0** | 38 (35%) | 4.21 (2.04, 8.71) | 0.0001 | 3.41 (1.55, 7.51) | 0.002 |
| **By 1 mmol/L Increase in CSF Lactate** |  |  |  |  |  |
| **1 unit increase** |  | 1.16 (1.07, 1.27) | 0.0005 | 1.13 (1.02, 1.24) | 0.02 |
| **By AUC Cut Off of CSF Lactate** |  |  |  |  |  |
| **<4.3** | 25 (14%) | 1.0 |  | 1.0 |  |
| **≥4.3** | 47 (33%) | 2.53 (1.56, 4.12) | 0.0002 | 2.10 (1.22, 3.62) | 0.008 |

\*Adjusted for Glasgow Coma Score, baseline seizure, quantitative cryptococcal CSF culture, and CSF opening pressures

**Table 3:** Baseline Characteristics of Patients with CSF Lactate >5.0 mmol/L, compared by 2-week survivor status.

| Characteristic | Died by 2 weeks | Survived 2 weeks | P-value |
| --- | --- | --- | --- |
| **N** | 38 | 71 |  |
| **Men** | 25 (66%) | 54 (76%) | 0.22 |
| **Age, years** | 40 [33, 50] | 37 [30, 42] | 0.06 |
| **Glasgow Coma Score < 15** | 29 (78%) | 42 (59%) | **0.05** |
| **Seizure** | 13 (35%) | 19 (27%) | 0.34 |
| **On HIV therapy** | 21 (55%) | 32 (46%) | 0.32 |
| **Hemoglobin, g/dL** | 10.2 [8.1, 12.6] | 11.6 [9.7, 13.1] | **0.04** |
| **CD4 count, cells/μL** | 20 [7, 52] | 27 [10, 70] | 0.33 |
| **Creatinine, mg/dL** | 0.8 [0.6, 1.7] | 0.8 [0.6, 0.9] | **0.001** |
| **CSF Lactate, mmol/L** | 7.0 [5.8, 7.7] | 6.7 [5.5, 8.5] | 0.57 |
| **CSF White cells, per μL** | 4.0 [4.0, 130.0] | 4.5 [4.0, 130.0] | 0.64 |
| **CSF Glucose, mg/dL** | 37.5 [20.0, 76.0] | 39.0 [26.0, 61.0] | 0.36 |
| **CSF Opening Pressure, cm H<sub>2</sub>O** | 27 [17, 43] | 26 [18, 37] | 0.22 |
| **CSF culture, log<sub>10</sub>CFU/mL** | 4.6 [2.6, 5.5] | 4.6 [2.8, 5.4] | 0.48 |
| **CSF removed at baseline, mL** | 13 [8, 20] | 12 [8, 18] | 0.63 |

Data are presented as median [IQR] or N (%)
P-values from Cox proportional hazards models

We assessed for other possible confounders for association with 2-week mortality, in those who presented with CSF lactate >5.0 mmol/L. There was a notable difference in the percentage of people with baseline Glasgow coma score <15 in those who died as compared to those who survived (died: 78% (29/38) vs. survived: 59% (42/71); p=.05). Among persons who died at 2-weeks, baseline hemoglobin was higher in those who survived (died: median 10.2 g/dL; IQR, 8.1 to 12.6 vs. survived: median 11.6 g/dL; IQR, 9.7 to 13.1; p=.04). There was also a difference in baseline serum creatinine levels, such that people who died at 2-weeks presented with higher baseline serum creatinine compared to those who survived, (p=.001). There were no significant differences in baseline CSF opening pressures (p=.22), CSF glucose (p=.36), or quantitative Cryptococcus CSF culture (p=.24) in persons who survived compared to those who died at 2-weeks.

Among persons with CSF lactate measurements at baseline, 37 individuals had matched capillary blood lactate also measured. No correlation existed between CSF and capillary blood lactate levels (r^2^=0.02; p=.60**) (Figure 3)**. Whole blood lactate levels did not differ between low, mid-range, and high CSF lactate tertiles (p=.72).

**Figure 3:**
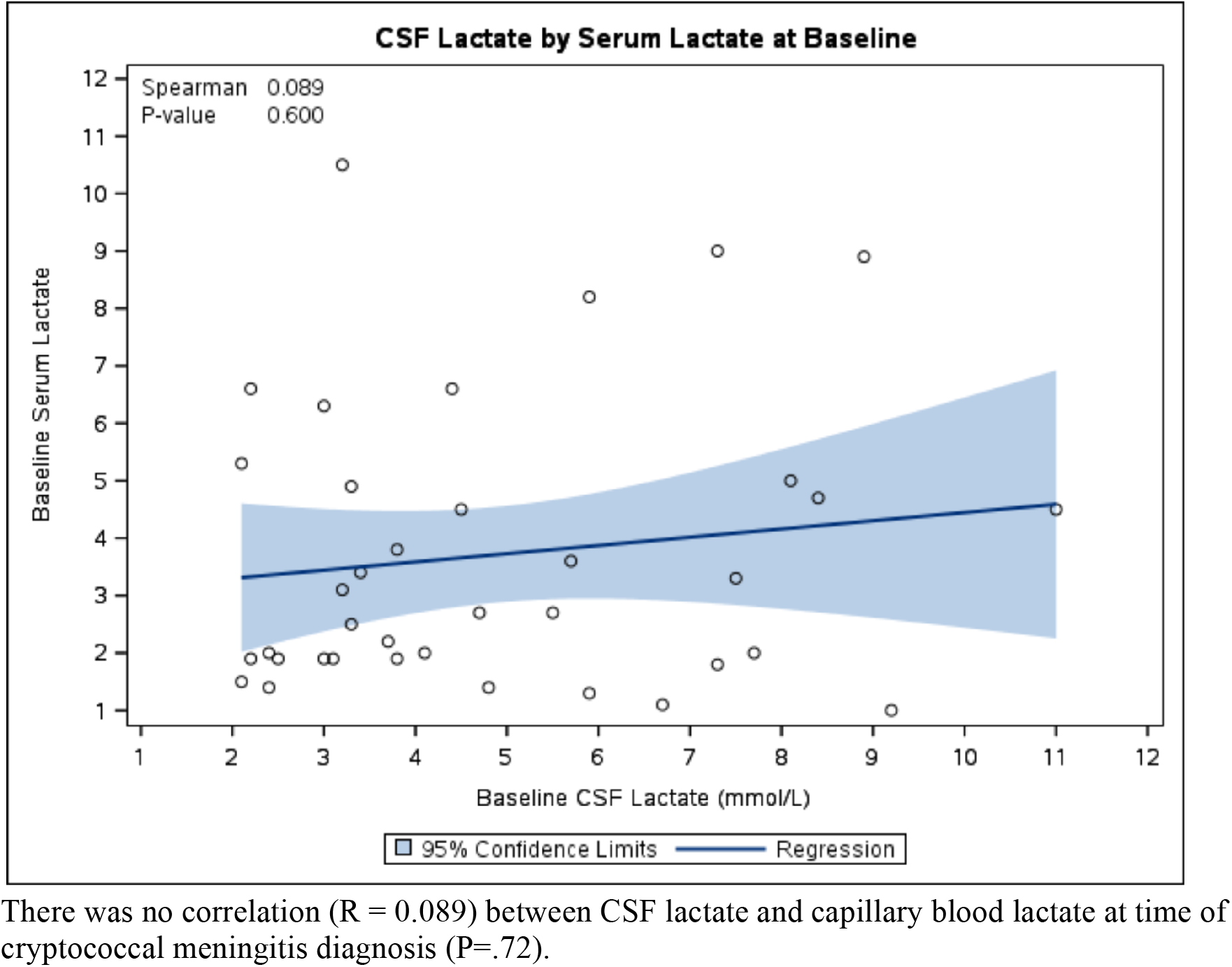
Relationship between CSF Lactate and Capillary Blood Lactate.

## Discussion

In this study, we demonstrate that a high baseline CSF lactate level is a reliable prognostic marker of disease severity in HIV-associated cryptococcal meningitis. Persons with HIV-associated cryptococcal meningitis who have elevated CSF lactate levels >5.0 mmol/L are also more likely to present with altered mental status (Glasgow coma score <15) and seizures at diagnosis. An elevated CSF lactate level >5.0 mmol/L is also accompanied by elevated intracranial pressures, higher CSF white cells, and lower CSF glucose. In contrast, CSF lactate levels are not associated with the burden of *Cryptococcus* based on a lack of association with quantitative CSF cultures. Persons with a baseline CSF lactate level >5.0 mmol/L were at a 3fold increased risk of death at 2-weeks compared to those with levels ≤3.0 mmol/L. Even after controlling for Glasgow coma score, opening pressure, and CSF quantitative *Cryptococcus* cultures, high CSF lactate (>5.0 mmol/L) was found to be an independent predictor for increased 2-week mortality. Assessing for clinical risk factors among persons with a CSF lactate of >5.0 mmol/L who died versus those who survived at 2-weeks, persons who died were more likely to also present with a lower hemoglobin value and higher creatinine levels at baseline.

While CSF lactate levels widely varied in cryptococcal meningitis, we did not find a correlation between CSF and capillary blood lactate measurements. Though lactate crosses the blood brain barrier through monocarboxylic acid transporters, the lack of correlation between CSF and capillary lactate levels may suggest a centrally occurring disease process, which is not systemic. Thereby suggesting that the increased mortality observed in persons with HIV-associated cryptococcal meningitis presenting with high baseline CSF lactate is a CNS driven process, which therefore requires compartment-specific interventions.

The exact mechanism behind CSF lactate elevations in cryptococcal meningitis and its association with mortality is unclear. We postulate that, in HIV-associated cryptococcal meningitis, decreases in cerebral perfusion and cerebral hypoxia leads to the clinical presentation of altered mental status, seizures, and increased intracranial pressures at the time of presentation. The observed CSF profile, increase in white cell count, low glucose, and high lactate would support an inflammatory CNS process that may be a result of or contributing to decreasing cerebral perfusion and cerebral hypoxia. Furthermore, when comparing 2-week survival among persons with cryptococcal meningitis who presented with a baseline CSF lactate >5.0 mmol/L, individuals who died had lower baseline hemoglobin levels. Our results would support the assumption that mortality may be driven by decreased cerebral perfusion and possibly cerebral ischemia.

In cryptococcal meningitis, percent regional cerebral tissue oxygenation (rSO_2_), as measured by cerebral oximetry, has been shown to be a predictor of 2-week mortality.[15] Such that individuals with low cerebral perfusion are >3 times more likely to die within 30-days. Similar to CSF lactate, cerebral oximetry is independently associated with mortality after having controlled for altered mental status, CSF quantitative *Cryptococcus* culture, CSF white cell count, and anemia. Low hemoglobin levels decrease oxygen carrying capacity and are strongly correlated with measured rSO_2_ and mortality. In fact, in previous cryptococcal meningitis cohorts, low hemoglobin levels have repeatedly been shown to be associated with increased mortality.[16-18]

While CSF glucose has long been recognized as highly variable in cryptococcal meningitis, the significance of glucose and other metabolic derangements has not been well characterized. The high-energy demand of the brain creates a state of dependence on aerobic metabolism and a continuously sufficient supply of oxygen and glucose for cellular metabolism. A lack of adequate cerebral perfusion, by either a reduction in the oxygen carrying capacity or cerebral ischemia secondary to increased intracranial pressures or seizures may explain the rise in CSF lactate. The oxidation of pyruvate and gluconeogenesis require the presence of oxygen. When oxygen levels fall, glucose is primarily converted to lactate. Thus, decreased cerebral perfusion leading to an increased risk for mortality would be supported by our observation of low CSF glucose levels in the presence of elevated CSF lactate.

High CSF lactate is independently associated with mortality, after controlling for increased intracranial pressures, altered mental status, and CSF quantitative *Cryptococcus* culture. This raises the possibility of irreversible CNS damage. In neurocritical care settings, cerebral microdialysis measurements of the lactate and pyruvate ratio help to distinguish between insufficient tissue oxygenation, which may be reversible and mitochondrial damage, which may lead to neural cell death.[19] Further research to assess changes in metabolite levels, in addition to lactate and pyruvate, in cryptococcal meningitis will be critical to understanding the degree of neurocellular damage in order to design targeted interventions to reduce mortality. Optimizing cerebral perfusion and oxygen delivery to the brain may be important aspects of the supportive management of neuroinfections.

While our study is the largest thus far in evaluating CSF lactate levels in cryptococcal meningitis, we were limited in our ability to longitudinally evaluate the effects of CSF lactate on outcomes. We were also limited by our sample size to focus on risk factors in individuals with high baseline CSF lactates >5.0 mmol/L. Despite these limitations, we believe that our study has important clinical implications in the management of cryptococcal meningitis.

In conclusion, results of our study demonstrate that in cryptococcal meningitis, baseline CSF lactate levels may be used as a prognostic marker for disease severity and as a clinical tool to risk stratify individuals who are at an increased risk of death. Our findings suggest that in individuals with cryptococcal meningitis, baseline CSF lactate levels >5.0 mmol/L should be used to identify those at increased risk of early, 2-week mortality to target further interventions. Further research needs to be done to better understand the underlying pathophysiology of elevated CSF lactate levels in cryptococcal meningitis.

## Data Availability

All data will be made available upon request.

